# Area-Level Economic Opportunity Modifies the Income-Health Gradient in the United States

**DOI:** 10.64898/2026.03.27.26349545

**Authors:** Anant Mishra, Rourke O’Brien, Atheendar Venkataramani

## Abstract

**Introduction:** Economic opportunity is a core pillar of the American Dream but is not distributed equally across communities. Substantial evidence has identified economic opportunity as an independent social determinant of health, but relatively little is known about opportunity’s relationship with other socioeconomic characteristics such as income. Here we address this gap in the literature to examine how area-level economic opportunity modifies the income-health gradient.

**Methods:** We used multivariable ordinary least squares models to estimate the association between self-reported health and economic opportunity across household income levels for working age adults (ages 25-64). Our measures of income and health come from the 2010-2019 Current Population Survey Annual Social and Economic Supplements. Our measure of economic opportunity was drawn from Opportunity Insights and represents the county-averaged national income percentile rank attained in adulthood for individuals born to parents at the 25^th^ percentile of the income distribution. We adjusted for a wide range of individual- and county-level demographic and socioeconomic characteristics.

**Results:** We find that county-level economic opportunity modified the gradient in self-reported health and household income among working-age adults. Effects were particularly pronounced in the lowest income deciles – an interdecile increase in economic opportunity was associated with closing almost 33% of the gap in health between the lowest and highest income deciles. The results were robust to sensitivity analyses.

**Conclusion:** We show that local area economic opportunity flattens the relationship between household income and health, with lower-income individuals benefitting the most from living in high opportunity areas.

## 1. Introduction

The American Dream rests on the promise of economic opportunity – the idea that an individual’s prospects for upward mobility are not determined by their family background. Yet, recent evidence suggests that economic opportunity is fading for many Americans, with economic outcomes of children from the poorest and wealthiest families widening over time (Chetty et al., 2026; Sawhill & McMurrer, 1996). Only 50% of Americans born in the 1980s could expect to attain a higher income than their parents compared to 90% for Americans born in the 1940s (Chetty et al., 2017). Administrative tax data reveal that economic opportunity is not distributed equally across race, sex, geography, or time (Chetty et al., 2016, 2018, 2020, 2026). Poor white children born in the 1990s had near universally worse social and economic outcomes compared to poor white children born a decade earlier (Chetty et al., 2024). And still, even Black children fortunate enough to grow up in counties with the highest levels of upward mobility ended up worse off, on average, in adulthood than white children from counties with the lowest levels of upward mobility. Further, economic opportunity is tightly stratified by educational attainment. For children who grow up in the poorest households, nearly 50% without a college degree remain in the lowest income group as adults compared to less than 20% with a college degree (Haskins, 2016).

A growing body of evidence has identified economic opportunity as an independent social determinant of health. Studies that examine the relationship between area-level intergenerational upward mobility and population health markers such as mortality rates find strong correlations that persist even after adjusting for other demographic, social, and economic characteristics (Gugushvili et al., 2025; O’Brien et al., 2020; Venkataramani, Brigell, et al., 2016; Venkataramani, Chatterjee, et al., 2016; Xiong et al., 2025; Zang & Tian, 2025). Natural experiment studies of policies or events that expand or restrict economic opportunities suggest that these associations reflect cause and effect (Finkelstein et al., 2026; Moorthy & Shaloka, 2025; O’Brien et al., 2022; Venkataramani et al., 2017, 2019, 2020). These relationships likely reflect multiple channels. Economic opportunity may contribute to improvements in known social determinants of health such as better employment, education, and access to healthcare. Economic opportunity may also alter the expected economic and social returns to health investments (Grossman, 1972). That is, in the absence of economic opportunity, the returns for investments in positive health behaviors are limited. Conversely, rising economic opportunity may induce health investments because the potential economic returns to engaging in healthy behaviors will increase (Venkataramani, Chatterjee, et al., 2016). Finally, recent literature has emphasized the role economic opportunity plays in conditioning one’s hopes and aspirations for a better future, especially for individuals living in poverty (Ray, 2006). Insights from psychology and economics suggest that hopes and aspirations may not only enhance intrinsic motivation to engage in healthy behaviors but also directly influence mental health (de Quidt & Haushofer, 2016). Lowered aspirations can reduce the motivation to engage in healthy behaviors and worsen mental health – for example, minority young adults had increased rates of tobacco and alcohol use following affirmative action bans compared to their non-minority peers (Case & Deaton, 2023; Venkataramani et al., 2019).

In addition to its direct association with health, economic opportunity may also modify relationships between socioeconomic characteristics, such as income, and health. The reasons for this follow from the same mechanisms directly linking opportunity and health. High opportunity areas may raise beliefs and aspirations of lower-income individuals in ways that improve health behaviors and outcomes. High opportunity areas also tend to have social networks that cross class boundaries, fostering peer effects and exposure to diverse information that may affect health behaviors and outcomes (Bor et al., 2024; Chetty et al., 2022; Eisenberg et al., 2014; Hinnosaar & Liu, 2022).

Despite this theoretical premise, there is no work to our knowledge that has examined how area-level economic opportunity modifies the income-health gradient. We address this gap in the literature using detailed individual survey data on working-age adults in the United States and high-resolution estimates of county-level economic opportunity.

## 2. Methods

### 2.1 Data Sources and Sample

Data on county-level economic opportunity were drawn from Opportunity Insights, “The Opportunity Atlas: Mapping the Childhood Roots of Social Mobility” (Chetty et al., 2018; Opportunity Insights, 2018b). For each county, we use as our primary measure of economic mobility the income percentile rank attained in adulthood (ages 31-37) for individuals born to parents at the 25^th^ percentile of the income distribution. Details regarding the precise construction of these estimates are described elsewhere. Briefly, federal tax return data for over 20 million children born between 1978-1983 were linked to their parents’ tax returns while growing up, 2000 and 2010 decennial Census data, and the 2005-2015 American Community Surveys in order to estimate children’s incomes in adulthood and their parents’ household income level (Chetty et al., 2018). Children were assigned to counties in proportion to the amount of their childhood they spent residing there. Of note, we subtracted the minimum values from the economic opportunity measure to facilitate interpretation.

Our measures of income and health come from 2010-2019 Current Population Survey Annual Social and Economic Supplements (CPS-ASEC). These data are publicly available from the Minnesota Population Center’s Integrated Public Use Microdata Project (IPUMS) (US Census Bureau, 2020). To preserve respondent confidentiality, only counties with 100,000 residents or more are identified in CPS-ASEC, which covers approximately 45% of the sample.

We restrict our sample to working age adults (age 25-64) as this is the population for whom economic opportunity is most relevant. However, we run separate sensitivity analyses for those age 65 or older. To allow for non-linear relationships between income and health, we transform household income into deciles to facilitate interpretation of the opportunity-health gradient across the income distribution (however, we include analyses using logarithmic transformations of household income in Appendix Table 5). We focus on the county-level because it is the smallest geographical unit for which the effects of opportunity are similar across individuals and for which there are detailed covariate data (see below).

### 2.2 Outcomes

Our primary outcome was a binary variable capturing the percentage of respondents who self-reported being in “good” or better health on a 5-point Likert scale: “Would you say that your health in general is excellent, very good, good, fair, or poor?” We also include additional sensitivity analysis treating respondents’ answers as a continuous measure.

### 2.3 Covariates

We adjust for several individual and county-level characteristics in our analyses. Individual-level demographic variables include age, sex, race (non-Hispanic white, black, Hispanic, other origin), and marital status. We also include binary indicators for high school and college completion, employment, and health insurance status.

For county-level socioeconomic covariates, we include county per capita income, unemployment rate, and Gini coefficient obtained from the US Bureau of Economic Analysis, US Bureau of Labor Statistics, and the US Census Bureau, respectively (US Bureau of Economic Analysis, 2015; US Bureau of Labor Statistics, 2020; US Census Bureau, 2015b). We also include demographic composition (the percentage of the population that was African American, the percentage of the population older than 65 years, and the percentage of the population 15 years or younger), population density, and a county’s urban-rural classification (counties in metropolitan areas, counties outside of metropolitan areas with an urban population >20,000, counties outside of metropolitan areas with an urban population between 2,500 and 20,000, and rural counties – populations <2,500). These data were drawn from the US Census Bureau and the US Department of Agriculture, respectively (US Census Bureau, 2015a, 2015c, 2018; Economic Research Service, US Department of Agriculture, 2013).

We also account for differences in county-level social and health structure. We include measures of county-level violent crime per capita based on Federal Bureau of Investigation Uniform Crime Statistics data available at the Inter-University Consortium for Political and Social Research (Federal Bureau of Investigation, 2019). We also include measures of social capital and connectedness drawn from Opportunity Insights “Social Capital I: Measurement and Associations with Economic Mobility” which captures the extent to which individuals of low-and high-socioeconomic status interact at the county-level (Chetty et al., 2022; Opportunity Insights, 2018a). We further include US Census-based measures of residential racial segregation obtained from the University of Wisconsin’s County Health Rankings (University of Wisconsin Public Health Institute, 2016). Finally, we include a measure of primary care physicians per capita drawn from US Health Resources and Services Administration data, but also available at the University of Wisconsin’s County Health Rankings (University of Wisconsin Public Health Institute, 2016).

Much of our county demographic data are drawn from a range of several years during our study period. For example, we average the county unemployment rate from 2010-2019 for each county and rely primarily on county-level estimates of age, race, and income spanning the first five years of our study period. However, for those measures where data across a time range was unavailable, we sought estimates as close to 2015 – the midpoint of our period of interest – as possible (e.g., urban-rural status, primary care physicians per capita). Inherent to this decision is the assumption that 2015 is a representative year for each county for these measures and that these measures are unlikely to have changed meaningfully across our study period. (Precise descriptions and data sources for all outcome, exposure, and covariate measures are provided in the Appendix Table 1.)

### 2.4 Statistical Analysis

For our analysis, we fitted multivariable ordinary least square (OLS) regression models of the following form:

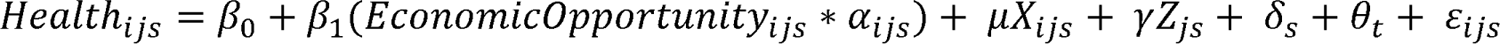

where *i* indexes individuals, *j* indexes counties, and *s* indexes the state of residence. *Health_ijs_* refers to the binary outcome variable measuring an individual’s general health status. (*EconomicOpportunity_js_ x* α*_ijs_)* represents the interaction term between the county-level measure of economic mobility and individual household income or educational attainment. The vectors *X_ijs_*and *Z_js_* are comprised of the individual-level and county-level covariates described previously. All models include state (δ*_s_*) and year (θ*_t_*) fixed effects to adjust for macrolevel socioeconomic and temporal factors that may be correlated with both economic opportunity and health. All standard errors are clustered at the county-level given that this is the level of variation for our exposure of interest.

Our main analysis consists of two sets of adjusted models. In the first, we include only a minimal set of individual covariates: household income, age, race, sex, and year fixed effects. In the second, we include the full set of individual- and county-level covariates described previously. We then assessed the stability of the coefficient estimates across these specifications, reasoning that stable coefficient estimates would suggest the robustness of the results to both omitted variable bias and overadjustment. We further apply these models to gauge the differential effect on sex and race.

In addition to these models, we conducted several sensitivity analyses. First, we included additional interaction terms between county-level demographic and social factors and household income to assess whether our estimates of the interaction between economic opportunity and income may reflect omitted interactions between other county-level characteristics and income.

In our second sensitivity analysis, we restrict the population of interest to only those who have not moved counties in the prior year to limit potential selection of healthier individuals into higher opportunity counties. Third, we switch our outcome variable from the binary indicator of good health to the original 5-point scale of health treated as an approximately continuous variable to gauge whether the effects seen are consistent across the health distribution. Lastly, we apply our model to individuals past retirement age (adults aged 65 and older) for which opportunity should theoretically be less meaningful.

All prevalence estimates and regression models were weighted by CPS-ASEC sample weights. All analyses were conducted using Stata, version 18 (StataCorp). This study relied solely on data in the public domain, so no ethical approval was sought for the study procedures.

## 3. Results

Our final sample consisted of more than 434,000 individuals between the ages of 25-64 during the CPS-ASEC survey period of 2010-2019. These individuals resided in 368 counties spanning 45 states. Table 1 summarizes both individual- and county-level characteristics for our entire sample population, as well as for those individuals residing in the bottom and top decile economic opportunity counties. Across our entire sample population, the mean age was 43.7 years and 43% reported being non-white. The median household income was reported as $65,503, 73% reported currently being employed, and 37% reported at least having a college degree. More than 83% reported having health insurance and just under 12% reported fair or poor health. The mean county-level unemployment rate during the study period was 6.5%. Approximately 1% of the sample resided in a rural county. The mean county-level economic opportunity value (after subtracting the minimum value) was 0.103 (range: 0.000 – 0.209).

**Table 1:**
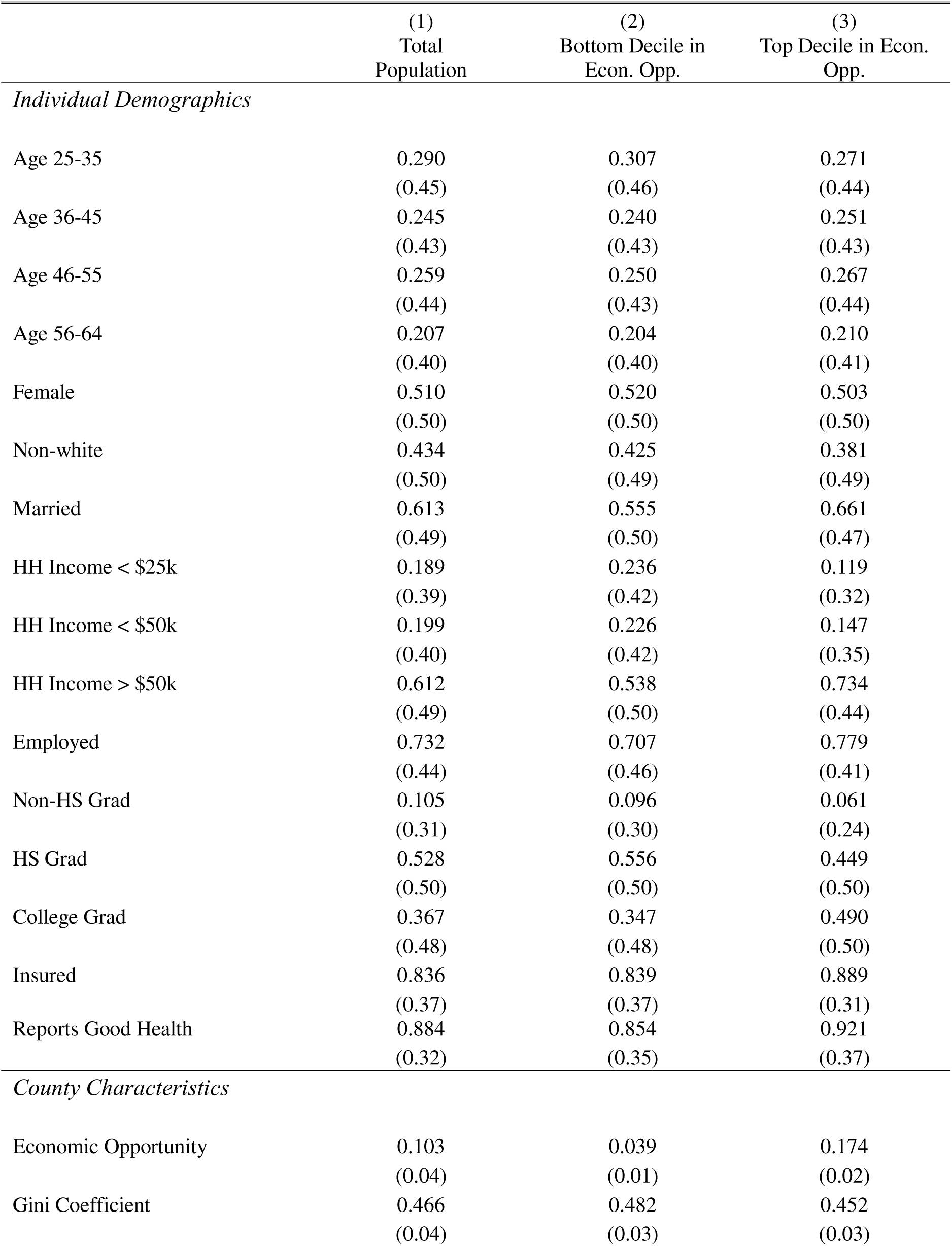
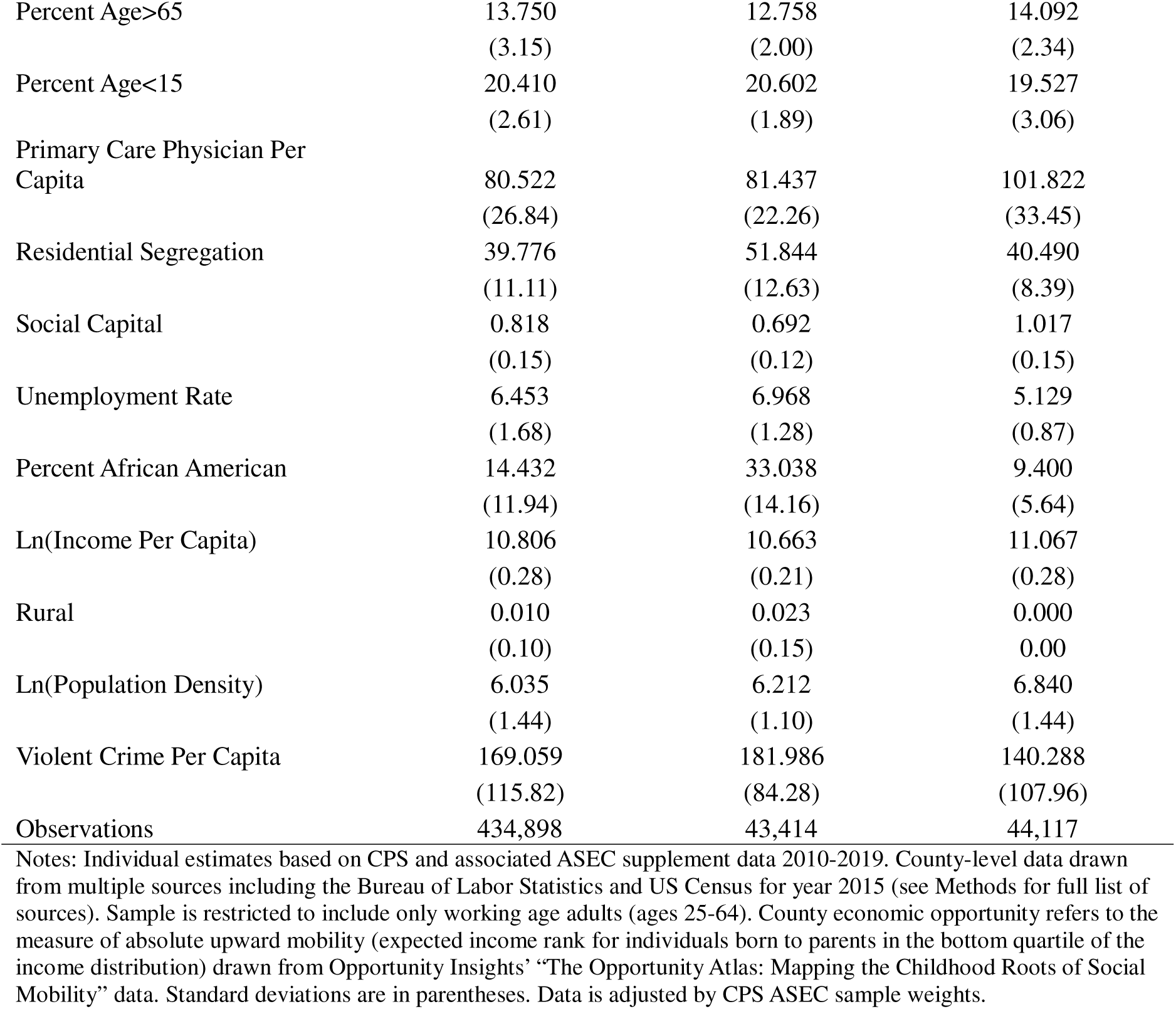
Demographics of the working age population (ages 25-64) and county characteristics for all individuals residing in and counties comprising the bottom and top decile counties for economic opportunity.

When comparing individuals residing in the bottom and top decile economic opportunity counties, individuals residing in the bottom decile of economic opportunity counties tended to be more non-white (43%) versus those in the top decile of economic opportunity (38%) and were less likely to be married (56% vs 66%). Those residing in the bottom decile of economic opportunity counties were also more likely to make less than $25,000 (24% vs 12%), and less likely to report being good or better health (85% vs 92%). Individuals living in the bottom decile of economic opportunity tended to reside in counties with higher degrees of residential segregation and less exposure to social capital. They also tended to reside in counties with a higher proportion of African Americans compared to the top decile of economic opportunity counties (33% vs 9%). The mean county-level unemployment rate for individuals living in the bottom decile of economic opportunity during the study period was 7% compared to 5% for individuals in the top decile of economic opportunity.

Higher economic opportunity was associated with improved probability of reporting good or better health across both high and low household income levels (Table 2; Figure 1). For example, in the baseline model with only a minimal set of individual covariates, an interdecile increase in economic opportunity was associated with a 1.1 percentage point increase in the probability of reporting good health (β=0.108, 95% CI, 0.019-0.197) for those in the highest decile of household income. We estimated larger associations after adjusting for full set of individual- and county-level covariates, including state fixed effects. In this model, an interdecile increase in economic opportunity was associated with a 1.9 percentage point increase in the probability of reporting good health (β=0.188, 95% CI, 0.038-0.339) for those in the highest decile of household income. However, the effects of opportunity were particularly pronounced in the lowest income deciles. For individuals in the lowest income decile (on average, an annual household income of $6,200), an interdecile increase in economic opportunity was associated with a 7.8 percentage point increase in the probability of reporting good health or almost 33% of the gap in the probability of reporting good health between the between the lowest and highest income deciles in the sample.

**Figure 1.**
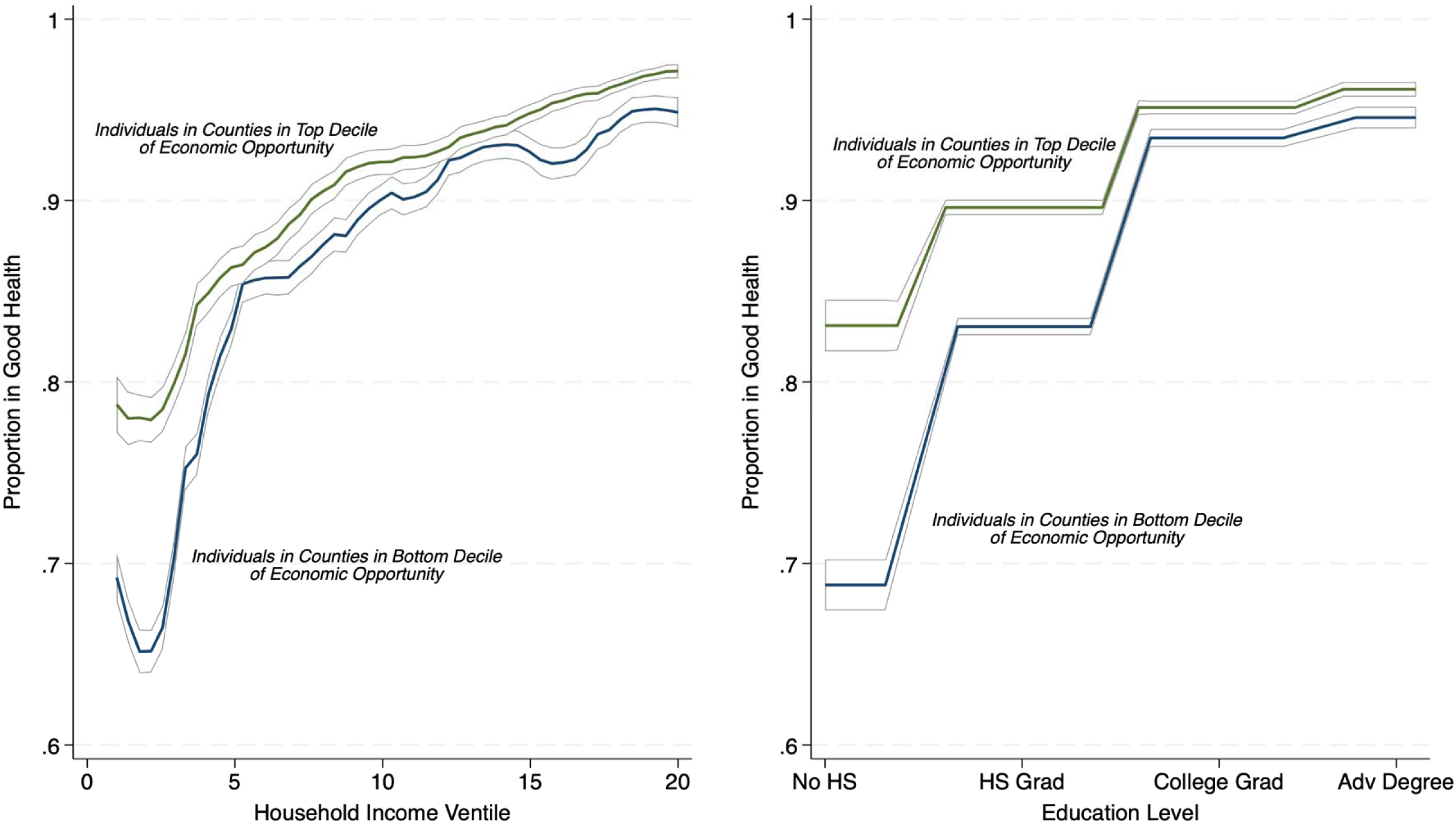
A and 1B. Income, education, health, and economic opportunity.

**Table 2:**
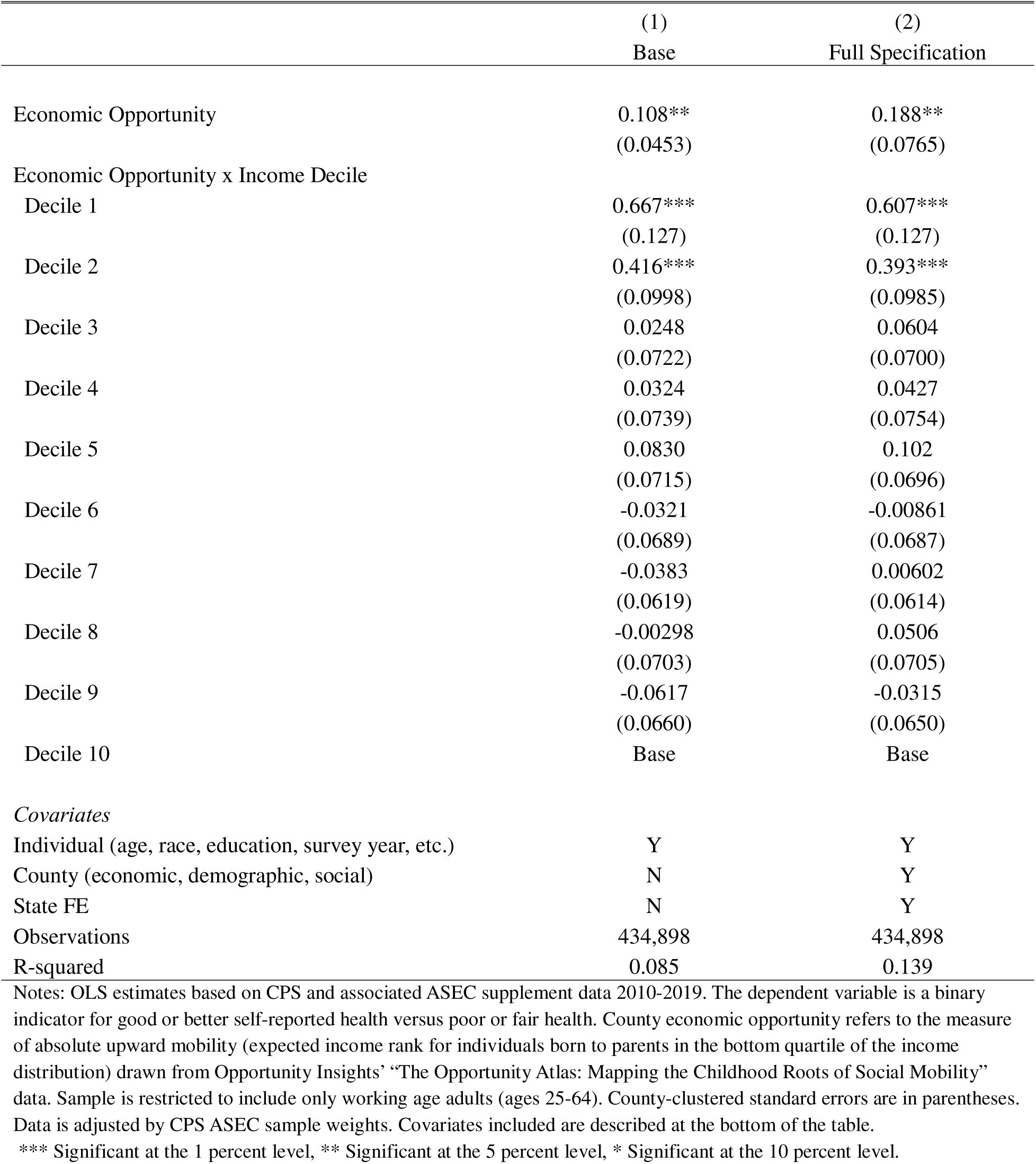
Economic opportunity as a moderator of the relationship between health and household income, particularly for low-income households.

This same positive association between the probability of reporting good health and higher economic opportunity was also seen across all levels of educational attainment, with particularly strong effect sizes for those with high school degrees or less than high school degrees (Table 3; Figure 1). For those with less than a high-school diploma, an interdecile increase in economic opportunity was associated with a 6.8 percentage point increase in the probability of reporting good health. This represents more than 40% of the gap in the probability of reporting good health between the between the least (no high school degree or equivalent) and most well-educated individuals (advanced degree) in the sample.

**Table 3:** Economic opportunity as a moderator of the relationship between health and educational attainment.

|  | (2)<br>Full Specification |
| --- | --- |
| Economic Opportunity | 0.154**<br>(0.0690) |
| Economic Opportunity x Educational Attainment |  |
| No High School Degree | 0.531***<br>(0.123) |
| High School or Equivalent Degree | 0.209***<br>(0.0514) |
| Bachelor's Degree | -0.0151<br>(0.0387) |
| Advanced Degree | Base |
| *Covariates* |  |
| Individual (age, race, income, survey year, etc.) | Y |
| County (economic, demographic, social) | Y |
| State FE | Y |
| Observations | 434,898 |
| R-squared | 0.139 |

Notes: OLS estimates based on CPS and associated ASEC supplement data 2010-2019. The dependent variable is a binary indicator for good or better self-reported health versus poor or fair health. County economic opportunity refers to the measure of absolute upward mobility (expected income rank for individuals born to parents in the bottom quartile of the income distribution) drawn from Opportunity Insights' "The Opportunity Atlas: Mapping the Childhood Roots of Social Mobility" data. Estimates reflect interaction between economic opportunity and self-reported educational attainment rather than income. Sample is restricted to include only working age adults (ages 25-64). County-clustered standard errors are in parentheses. Data is adjusted by CPS ASEC sample weights. Covariates included are described at the bottom of the table.
\*\*\* Significant at the 1 percent level, \*\* Significant at the 5 percent level, \* Significant at the 10 percent level.

We also find heterogenous effects across age, sex, and race. Economic opportunity is positively associated with the probability of reporting good health across household income for all age deciles in our sample but is particularly strong (and statistically significant) for individuals aged 25-35 and 46-55 (Appendix Table 3). However, the association of opportunity on health for the lowest income deciles was strongly positive across all age ranges. Economic opportunity had slightly stronger associations on the probability of reporting good health in women than men for both the highest and lowest income deciles (Appendix Table 4). When stratified by race, the effects of opportunity had significantly stronger associations for black and Hispanic individuals in the highest income decile compared to non-Hispanic whites (Appendix Table 5). Although all three race groups also exhibited sizable effects in the lowest income deciles, these associations reached statistical significance only among non-Hispanic whites.

We ran multiple sensitivity analyses to confirm the robustness of our findings. First, we show that our pattern of results remains robust to the inclusion of a large number of individual- and county-level interaction terms, including the inclusion of interactions between household income and county-level socioeconomic factors (Appendix Table 2, Column 3). In addition, our results remain largely unchanged when we restrict the population of interest to only those who have not moved counties in the prior year (Appendix Table 2, Column 4). We also observe similar magnitude in effect when we change our outcome variable to the original 5-point scale of health treated as an approximately continuous variable (Appendix Table 2, Column 5). Lastly, when we apply our model to those older than aged 65 for whom opportunity should theoretically be less salient, we observe diminished and non-significant effects compared to the working age population across both high and low-income levels (Appendix Table 3, Column 5).

## 4. Discussion

In this study, we find that county-level economic opportunity modified the gradient in self-reported health and household income among working-age adults across the US during the period 2010-2019. Higher opportunity areas both had flatter curves and higher intercepts (i.e., higher levels of health at all income levels) compared to lower opportunity areas. This finding persisted even after adjustment for multiple individual- and county-level characteristics and were robust to a number of different sensitivity tests.

These findings bolster the now substantial body of literature highlighting the association between local area economic opportunity and population health (Finkelstein et al., 2026; Gugushvili et al., 2025; Ludwig et al., 2011, 2012; Moorthy & Shaloka, 2025; O’Brien et al., 2020, 2022; Venkataramani, Brigell, et al., 2016; Venkataramani, Chatterjee, et al., 2016; Venkataramani et al., 2017, 2019, 2020; Xiong et al., 2025; Zang & Tian, 2025). We show that economic opportunity is positively associated with self-reported health for a broad contingent of working age adults across a large and relatively recent time period. Most notably, however, our findings advance this literature by characterizing the relationship between local area economic opportunity, health, and income. While the association between economic opportunity and self-reported health was meaningful and statistically significant across all household income levels, it was particularly magnified for respondents with the lowest levels of household income: an interdecile increase in economic opportunity for these individuals was equivalent to closing almost 33% of the gap in the probability of reporting good health between them and the highest income decile. Similar strong associations were seen between opportunity and educational attainment, especially for those with less than a high school diploma. Stated another way, local economic opportunity seems to matter most for the health of the most vulnerable members of society.

Our findings have important implications for policymakers. First, they reinforce the idea that policies aimed at improving local economic opportunity may have meaningful consequences for population health, and that interventions aiming to improve population health must consider the role expanding economic opportunity can play, especially for vulnerable populations. Our results also emphasize the outsize influence that places and their surrounding opportunity structures exert on health. In particular, the finding that economic opportunity moderates the income-health gradient suggests that improving opportunity may attenuate health disparities even in the absence of large changes in household income. In essence, augmenting opportunity – such as through improving labor market access, education quality, or infrastructure – may partially buffer the adverse health effects of lower socioeconomic status. This may be especially relevant in those localities with the lowest levels of economic opportunity where opportunity investments may yield disproportionate health benefits.

These results should be interpreted in light of several limitations. First, while we adjust for a large number of individual- and county-level covariates and include a falsification test for those aged 65 and older, we cannot exclude that our findings might have been affected by omitted variables; that is, our results should be interpreted as correlational but not causal. Second, although we did not observe meaningful changes in our findings when restricting our analyses to those individuals who had not moved counties in the prior year, bias from non-random residential migration of healthier individuals to high opportunity areas within counties or over a longer period cannot be excluded. Third, our measure of economic opportunity from Opportunity Insights was retrospective and based on outcomes from individuals born between 1978 and 1983. County-level economic opportunity during our study period may have differed from these estimates, though analyses have shown that these prior estimates are persistent to the present (Chetty et al., 2018). More broadly, our estimates are subject to the limitations of the underlying economic mobility data from Opportunity Insights, though they remain the most detailed and comprehensive data available on geographic economic mobility to date (Chetty et al., 2018).

Fourth, we focus on the county-level in part due to data availability, but economic opportunity and population health have been shown to have salient effects at a more granular level (such as the census tract or neighborhood) (Boing et al., 2020; Chetty et al., 2018). Furthermore, CPS-ASEC data is only available for counties with populations of approximately 100,000 or more – it is possible that our findings may differ in less-populated counties. We also draw on data for many of our county-level covariates that span most but not all of our period of interest, though we suspect these data did not change meaningfully in years not explicitly captured. Finally, our primary health measure was self-reported and therefore prone to reporting bias.

## 5. Conclusion

We show that local area economic opportunity flattens relationship between household income and health, with lower-income individuals benefitting the most from living in high opportunity areas. Local, state, and national policymakers can use these findings as further evidence that policies aimed at expanding economic opportunity may play an important role in improving population health.

## Author Contributions

**Anant Mishra:** Writing – original draft; Writing – review and editing; Data curation; Formal analysis; Methodology; Conceptualization. **Rourke O’Brien:** Writing – review and editing; Supervision; Project Administration; Methodology; Conceptualization. **Atheendar Venkataramani:** Writing – review and editing; Supervision; Project Administration; Funding acquisition; Methodology; Conceptualization.

## Data Availability

https://cps.ipums.org/cps/

Links to all data used are cited within the manuscript. Further description of the data sources can be found in Supplementary Appendix Table 1.

## Appendix Tables and Figures

**Appendix Table 1:**
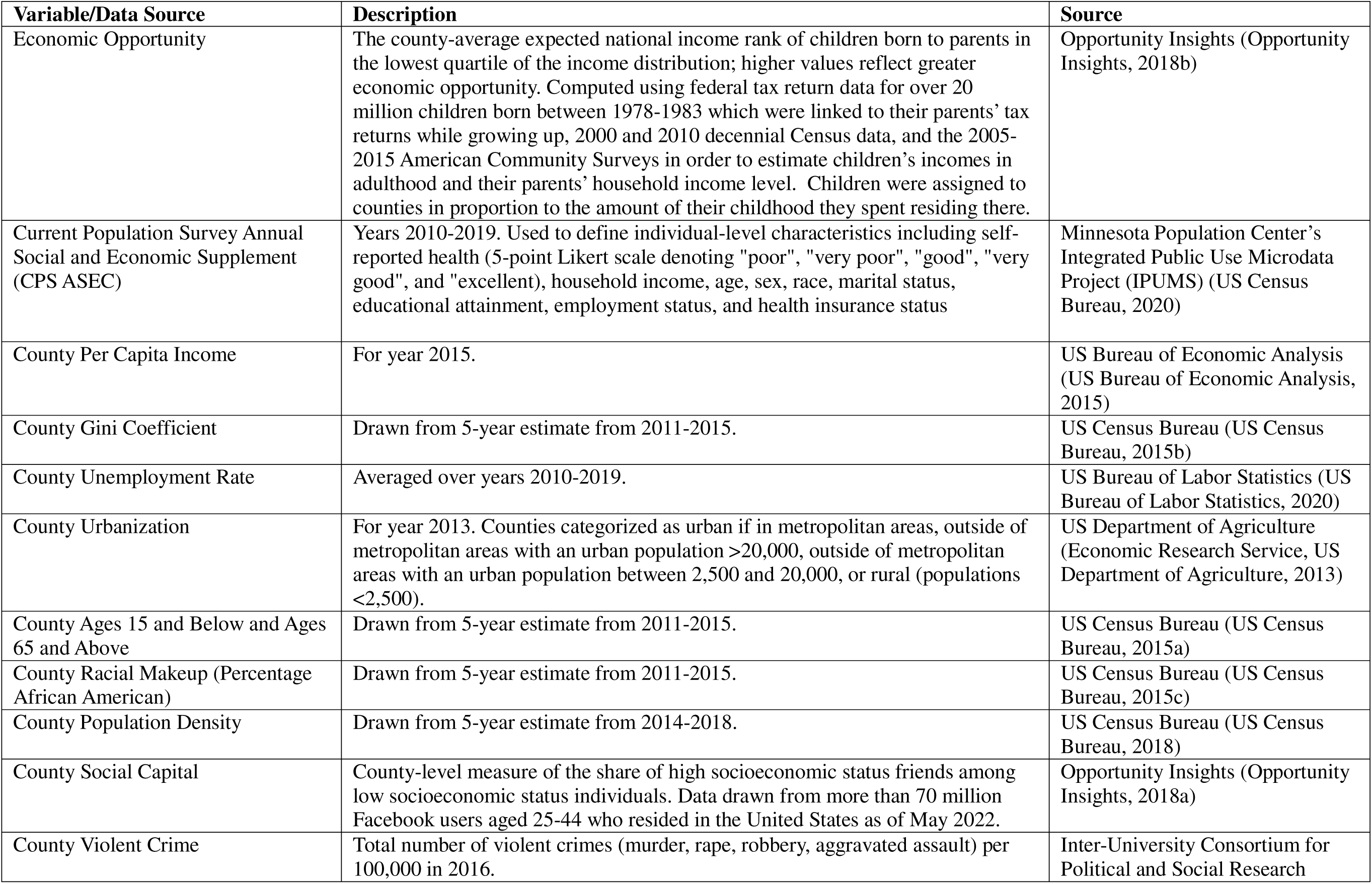
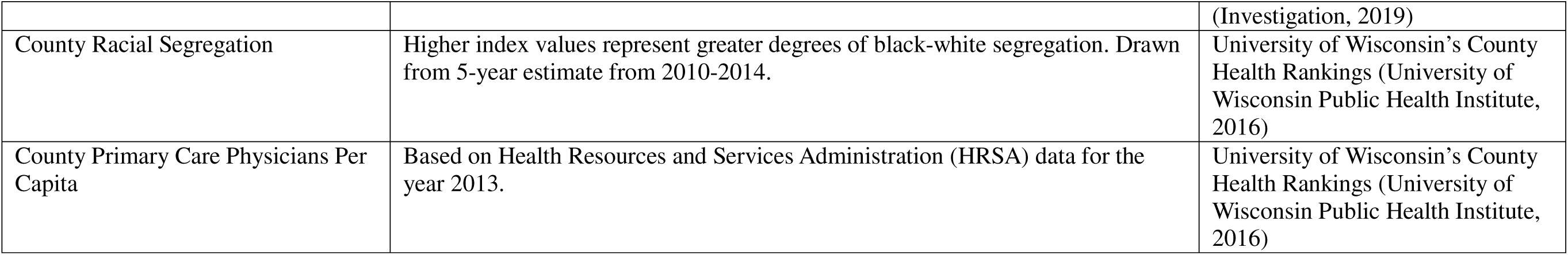
Variable and Data Sources Summary.

| Variable/Data Source | Description | Source |
| --- | --- | --- |
| Economic Opportunity | The county-average expected national income rank of children born to parents in the lowest quartile of the income distribution; higher values reflect greater economic opportunity. Computed using federal tax return data for over 20 million children born between 1978-1983 which were linked to their parents' tax returns while growing up, 2000 and 2010 decennial Census data, and the 2005-2015 American Community Surveys in order to estimate children's incomes in adulthood and their parents' household income level. Children were assigned to counties in proportion to the amount of their childhood they spent residing there. | Opportunity Insights (Opportunity Insights, 2018b) |
| Current Population Survey Annual Social and Economic Supplement (CPS ASEC) | Years 2010-2019. Used to define individual-level characteristics including self-reported health (5-point Likert scale denoting "poor", "very poor", "good", "very good", and "excellent), household income, age, sex, race, marital status, educational attainment, employment status, and health insurance status | Minnesota Population Center's Integrated Public Use Microdata Project (IPUMS) (US Census Bureau, 2020) |
| County Per Capita Income | For year 2015. | US Bureau of Economic Analysis (US Bureau of Economic Analysis, 2015) |
| County Gini Coefficient | Drawn from 5-year estimate from 2011-2015. | US Census Bureau (US Census Bureau, 2015b) |
| County Unemployment Rate | Averaged over years 2010-2019. | US Bureau of Labor Statistics (US Bureau of Labor Statistics, 2020) |
| County Urbanization | For year 2013. Counties categorized as urban if in metropolitan areas, outside of metropolitan areas with an urban population >20,000, outside of metropolitan areas with an urban population between 2,500 and 20,000, or rural (populations <2,500). | US Department of Agriculture (Economic Research Service, US Department of Agriculture, 2013) |
| County Ages 15 and Below and Ages 65 and Above | Drawn from 5-year estimate from 2011-2015. | US Census Bureau (US Census Bureau, 2015a) |
| County Racial Makeup (Percentage African American) | Drawn from 5-year estimate from 2011-2015. | US Census Bureau (US Census Bureau, 2015c) |
| County Population Density | Drawn from 5-year estimate from 2014-2018. | US Census Bureau (US Census Bureau, 2018) |
| County Social Capital | County-level measure of the share of high socioeconomic status friends among low socioeconomic status individuals. Data drawn from more than 70 million Facebook users aged 25-44 who resided in the United States as of May 2022. | Opportunity Insights (Opportunity Insights, 2018a) |
| County Violent Crime | Total number of violent crimes (murder, rape, robbery, aggravated assault) per 100,000 in 2016. | Inter-University Consortium for Political and Social Research |
|  |  | (Investigation, 2019) |
| County Racial Segregation | Higher index values represent greater degrees of black-white segregation. Drawn from 5-year estimate from 2010-2014. | University of Wisconsin's County Health Rankings (University of Wisconsin Public Health Institute, 2016) |
| County Primary Care Physicians Per Capita | Based on Health Resources and Services Administration (HRSA) data for the year 2013. | University of Wisconsin's County Health Rankings (University of Wisconsin Public Health Institute, 2016) |

---

**Appendix Table 2:**
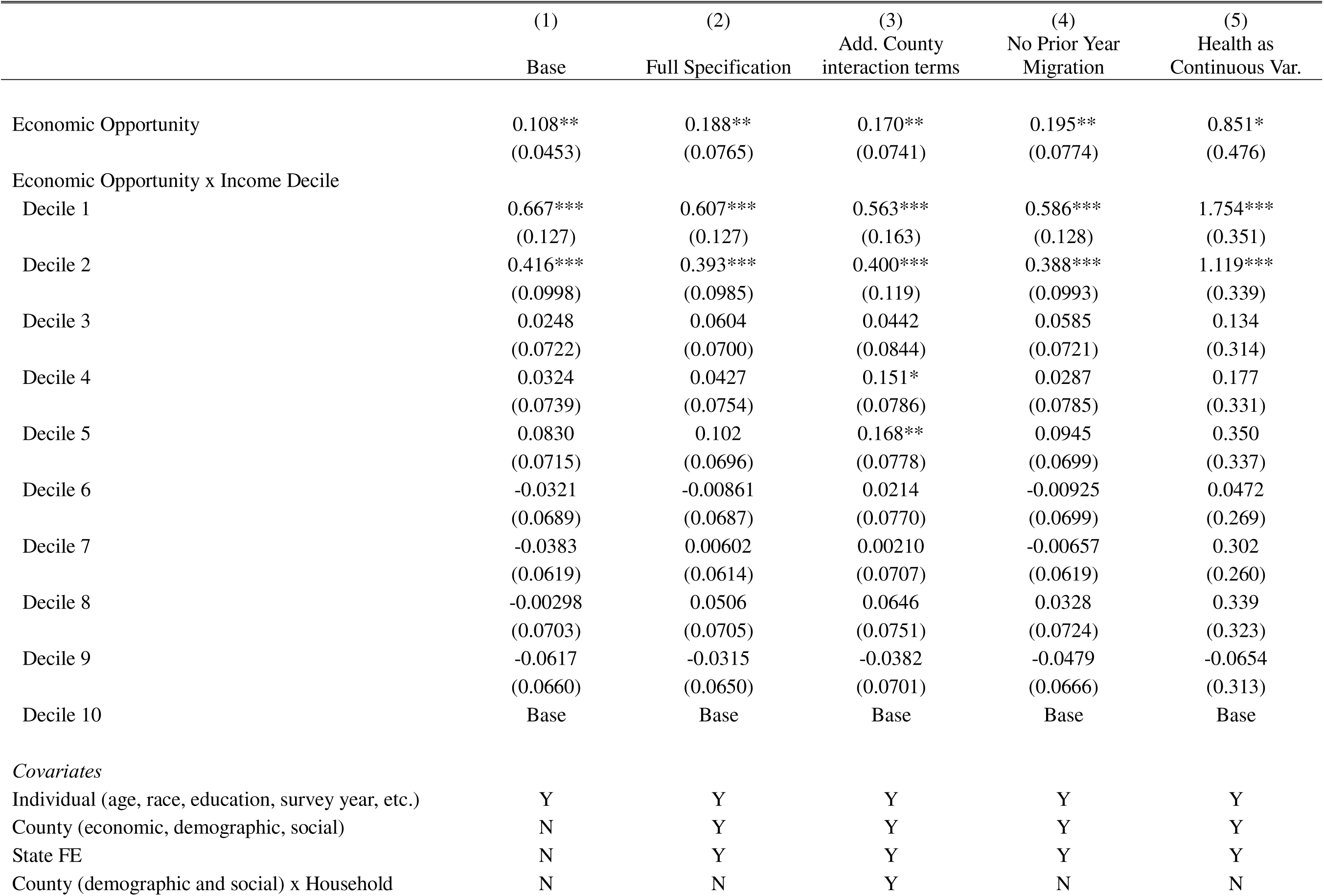
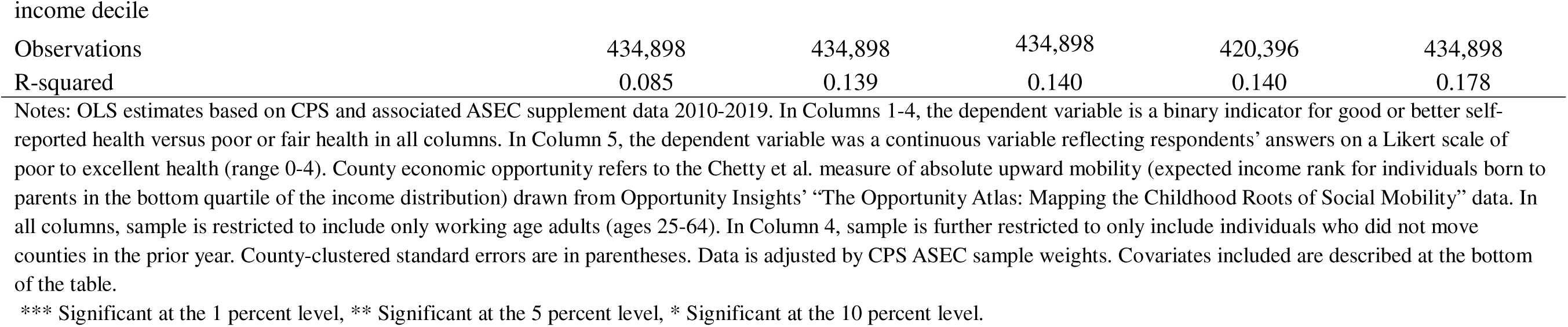
Economic opportunity as a moderator of the relationship between health and household income, particularly for low-income households.

|  | (1)<br>Base | (2)<br>Full Specification | (3)<br>Add. County<br>interaction terms | (4)<br>No Prior Year<br>Migration | (5)<br>Health as<br>Continuous Var. |
| --- | --- | --- | --- | --- | --- |
| Economic Opportunity | 0.108**<br>(0.0453) | 0.188**<br>(0.0765) | 0.170**<br>(0.0741) | 0.195**<br>(0.0774) | 0.851*<br>(0.476) |
| Economic Opportunity x Income Decile |  |  |  |  |  |
| Decile 1 | 0.667***<br>(0.127) | 0.607***<br>(0.127) | 0.563***<br>(0.163) | 0.586***<br>(0.128) | 1.754***<br>(0.351) |
| Decile 2 | 0.416***<br>(0.0998) | 0.393***<br>(0.0985) | 0.400***<br>(0.119) | 0.388***<br>(0.0993) | 1.119***<br>(0.339) |
| Decile 3 | 0.0248<br>(0.0722) | 0.0604<br>(0.0700) | 0.0442<br>(0.0844) | 0.0585<br>(0.0721) | 0.134<br>(0.314) |
| Decile 4 | 0.0324<br>(0.0739) | 0.0427<br>(0.0754) | 0.151*<br>(0.0786) | 0.0287<br>(0.0785) | 0.177<br>(0.331) |
| Decile 5 | 0.0830<br>(0.0715) | 0.102<br>(0.0696) | 0.168**<br>(0.0778) | 0.0945<br>(0.0699) | 0.350<br>(0.337) |
| Decile 6 | -0.0321<br>(0.0689) | -0.00861<br>(0.0687) | 0.0214<br>(0.0770) | -0.00925<br>(0.0699) | 0.0472<br>(0.269) |
| Decile 7 | -0.0383<br>(0.0619) | 0.00602<br>(0.0614) | 0.00210<br>(0.0707) | -0.00657<br>(0.0619) | 0.302<br>(0.260) |
| Decile 8 | -0.00298<br>(0.0703) | 0.0506<br>(0.0705) | 0.0646<br>(0.0751) | 0.0328<br>(0.0724) | 0.339<br>(0.323) |
| Decile 9 | -0.0617<br>(0.0660) | -0.0315<br>(0.0650) | -0.0382<br>(0.0701) | -0.0479<br>(0.0666) | -0.0654<br>(0.313) |
| Decile 10 | Base | Base | Base | Base | Base |
| *Covariates* |  |  |  |  |  |
| Individual (age, race, education, survey year, etc.) | Y | Y | Y | Y | Y |
| County (economic, demographic, social) | N | Y | Y | Y | Y |
| State FE | N | Y | Y | Y | Y |
| County (demographic and social) x Household | N | N | Y | N | N |
| Observations | 434,898 | 434,898 | 434,898 | 420,396 | 434,898 |
| R-squared | 0.085 | 0.139 | 0.140 | 0.140 | 0.178 |

Notes: OLS estimates based on CPS and associated ASEC supplement data 2010-2019. In Columns 1-4, the dependent variable is a binary indicator for good or better self-reported health versus poor or fair health in all columns. In Column 5, the dependent variable was a continuous variable reflecting respondents' answers on a Likert scale of poor to excellent health (range 0-4). County economic opportunity refers to the Chetty et al. measure of absolute upward mobility (expected income rank for individuals born to parents in the bottom quartile of the income distribution) drawn from Opportunity Insights' "The Opportunity Atlas: Mapping the Childhood Roots of Social Mobility" data. In all columns, sample is restricted to include only working age adults (ages 25-64). In Column 4, sample is further restricted to only include individuals who did not move counties in the prior year. County-clustered standard errors are in parentheses. Data is adjusted by CPS ASEC sample weights. Covariates included are described at the bottom of the table.
\*\*\* Significant at the 1 percent level, \*\* Significant at the 5 percent level, \* Significant at the 10 percent level.

**Appendix Table 3:**
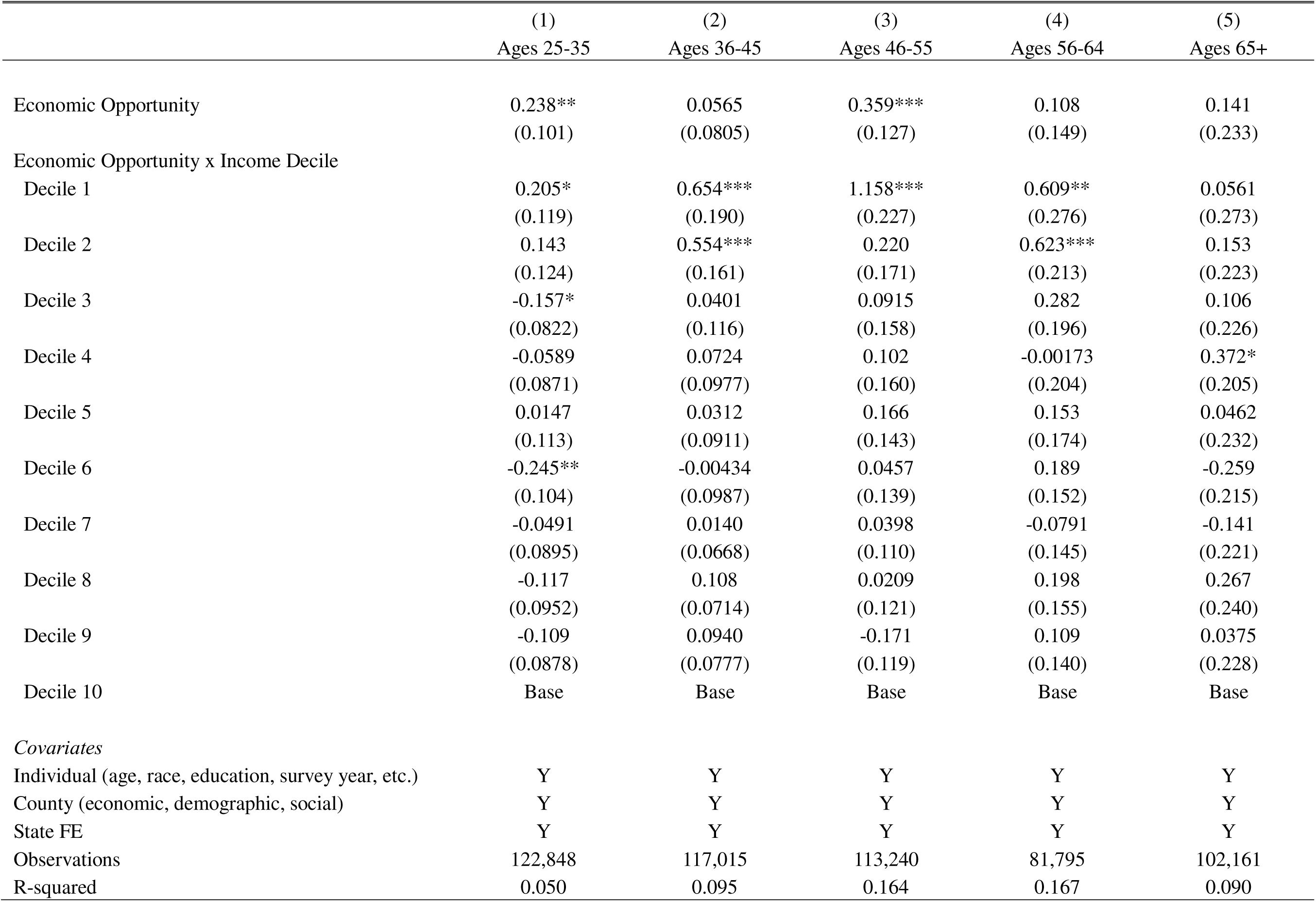
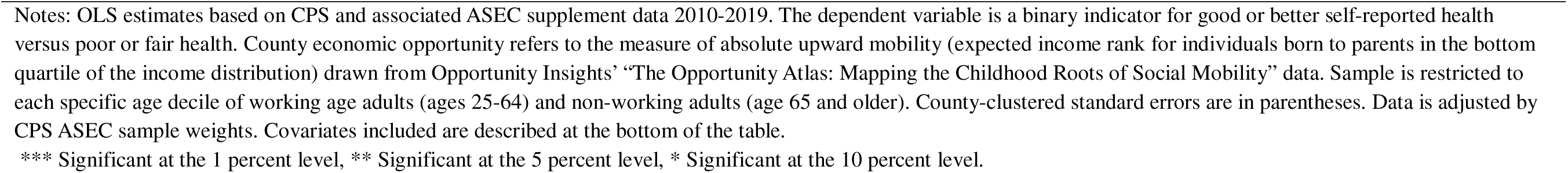
Economic opportunity as a moderator of the relationship between health and household income across working age (ages 25-64) and retiree (ages 65-up) population.

**Appendix Table 4:**
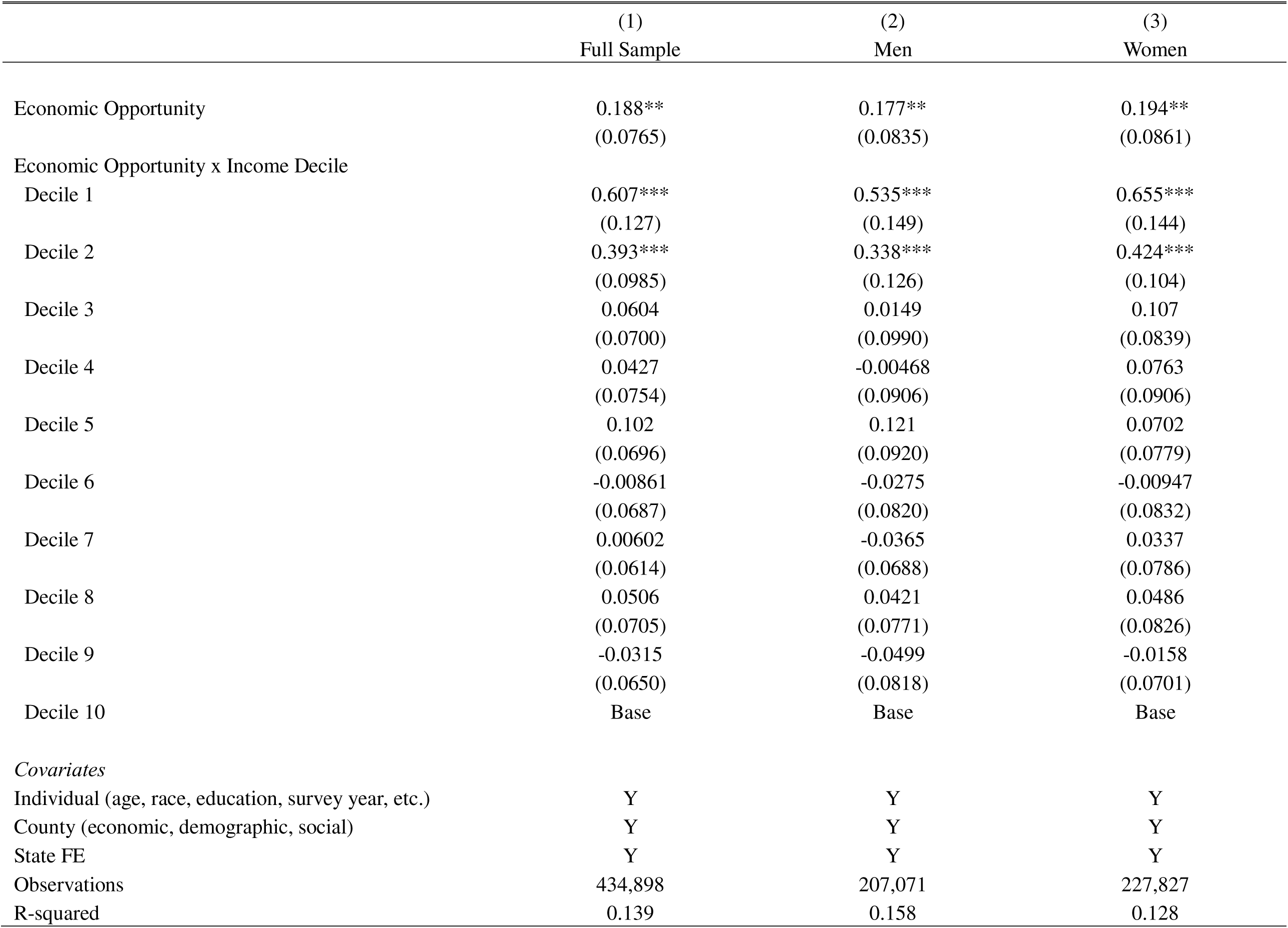
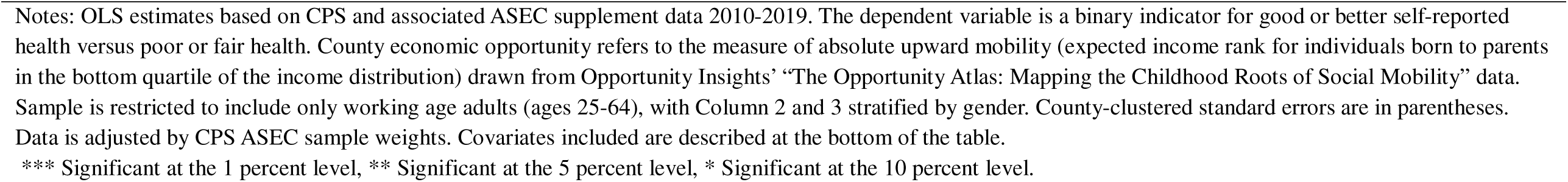
Economic opportunity as a moderator of the relationship between health and household income by gender.

**Appendix Table 5:**
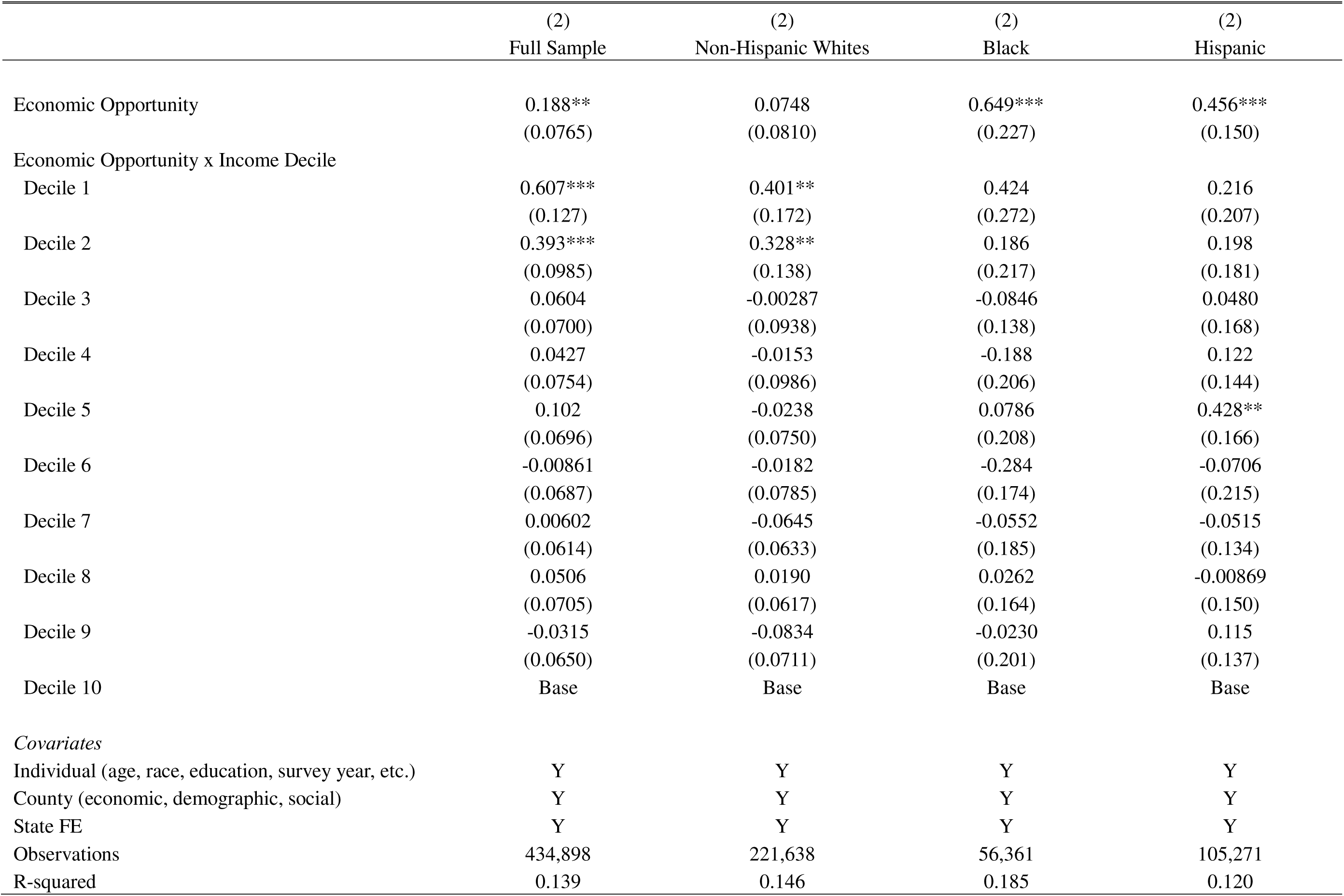
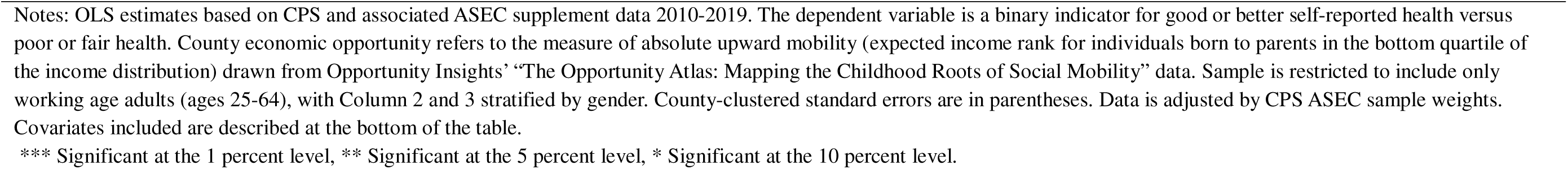
Economic opportunity as a moderator of the relationship between health and household income by race.

**Appendix Table 6:**
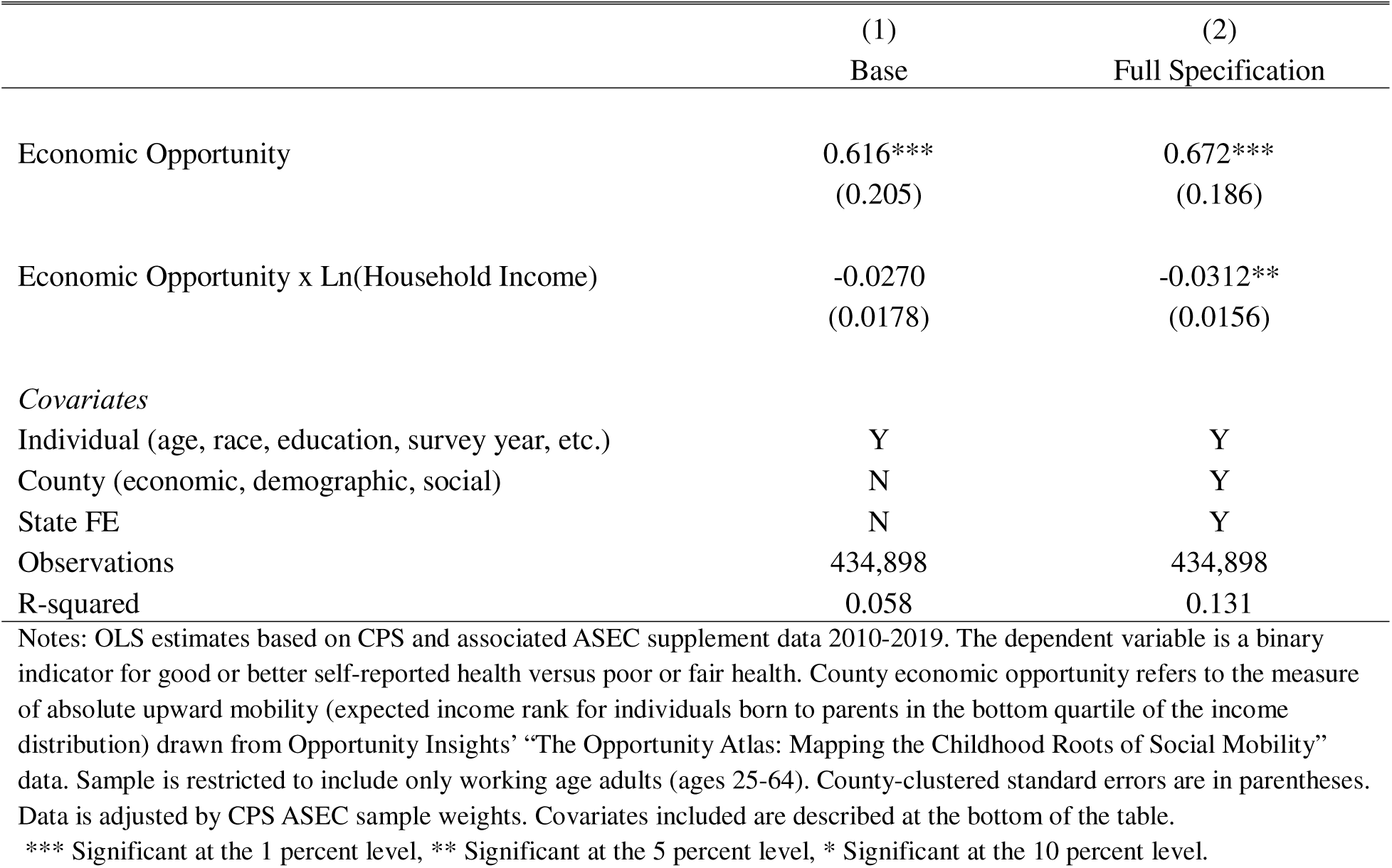
Economic opportunity as a moderator of the relationship between health and household income, particularly for low-income households using log-income.

